# European Autism GEnomics Registry (EAGER): Protocol for a multicentre cohort study and registry

**DOI:** 10.1101/2023.10.10.23296834

**Authors:** M. Bloomfield, A. Lautarescu, S. Heraty, S. Douglas, P. Violland, R. Plas, A. Ghosh, K. Van den Bosch, E. Eaton, M. Absoud, R. Battini, A. Blázquez Hinojosa, N. Bolshakova, S. Bolte, P. Bonanni, J. Borg, S. Calderoni, R. Calvo Escalona, M. Castelo-Branco, J. Castro-Fornieles, P. Caro, A. Danieli, R. Delorme, M. Elia, M. Hempel, N. Madeira, G. McAlonan, R. Milone, C. J. Molloy, S. Mouga, V. Montiel, A. Pina Rodrigues, C. P. Schaaf, M. Serrano, K. Tammimies, C. Tye, F. Vigevano, G. Oliveira, B. Mazzone, C. O’Neill, V. Romero, J. Tillmann, B. Oakley, D. Murphy, L. Gallagher, T. Bourgeron, C. Chatham, T. Charman

**Affiliations:** Department of Psychology, Institute of Psychiatry, Psychology & Neuroscience, King’s College London, London, United Kingdom; Department of Psychological Sciences, University of London, Birkbeck, London, United Kingdom; AIMS-2-Trials A-Reps, University of Cambridge, Cambridge, United Kingdom; Autism Research Centre, University of Cambridge, Cambridge, United Kingdom; Department of Children’s Neurosciences, Evelina London Children’s Hospital, Guy’s and St Thomas’ NHS Foundation Trust, London, United Kingdom; Department of Women and Children’s Health, Faculty of Life Sciences and Medicine, School of Life Course Sciences, King’s College London, London, United Kingdom; Department of Developmental Neuroscience, IRCCS, Fondazione Stella Maris, Pisa, Italy; Department of Clinical and Experimental Medicine, University of Pisa, Pisa, Italy; Department of Child and Adolescent Psychiatry and Psychology, Institute of Neurosciences, Hospital Clínic Universitari, Barcelona, Spain; Department of Psychiatry, School of Medicine, Trinity College Dublin, Dublin, Ireland; Center of Neurodevelopmental Disorders (KIND), Department of Women’s and Children’s Health, Centre for Psychiatry Research, Karolinska Institutet & Region Stockholm, Stockholm, Sweden; Child and Adolescent Psychiatry, Stockholm Health Care Services, Region Stockholm, Stockholm, Sweden; Curtin Autism Research Group, Curtin School of Allied Health, Curtin University, Perth, Australia; Epilepsy Unit, Scientific Institute IRCCS E. Medea Conegliano, Treviso, Italy; Centre for Psychiatry Research and Centre for Cognitive and Computational Neuropsychiatry (CCNP), Department of Clinical Neuroscience, Karolinska Institutet & Stockholm Health Care Services, Region Stockholm, Sweden; Psykiatri Affektiva, Sahlgrenska University Hospital, Gothenburg, Sweden; Institut d’Investigacions Biomèdiques August Pi i Sunyer (IDIBAPS), Barcelona, Spain; Centro de Investigación Biomédica en Red de Salud Mental (CIBERSAM), Spain; Department of Medicine, Institute of Neuroscience, University of Barcelona, Barcelona, Spain; CIBIT, ICNAS, Faculty of Medicine, University of Coimbra, Coimbra, Portugal; Institute of Human Genetics, University Hospital Heidelberg, Heidelberg, Germany; Child and Adolescent Psychiatry Department, Robert Debre Hospital, APHP, Université Paris Cité, Paris, France; Unit of Neurology and Clinical Neurophysiopathology, Oasi Research Institute-IRCCS, Troina, Italy; Psychiatry Department, Centro Hospitalar e Universitário de Coimbra, Coimbra, Portugal; Coimbra Institute for Biomedical Imaging and Translational Research, University of Coimbra, Coimbra, Portugal; Institute of Psychological Medicine, Faculty of Medicine, University of Coimbra, Coimbra, Portugal; Department of Forensic and Neurodevelopmental Sciences, Institute of Psychiatry, Psychology & Neuroscience, King’s College London, London, United Kingdom; Behavioural and Developmental Clinical Academic Group, South London and Maudsley NHS Foundation Trust, London, United Kingdom; CIBIT - Coimbra Institute for Biomedical Imaging and Translational Research, University of Coimbra, Coimbra, Portugal; ICNAS - Institute of Nuclear Sciences Applied to Health, University of Coimbra, Coimbra, Portugal; Pediatric Neurology Department, Hospital Sant Joan de Déu, Institut de Recerca Sant Joan de Déu, Esplugues de Llobregat, Barcelona, Spain; Astrid Lindgren Children’s Hospital, Karolinska University Hospital, Region Stockholm, Stockholm, Sweden; Neurological Sciences and Rehabilitation Medicine Scientific Area - Bambino Gesù Children’s Hospital, Rome, Italy; University Clinic of Pediatrics, Faculty of Medicine, University of Coimbra, 3000-602 Coimbra, Portugal; Child Developmental Center and Research and Clinical Training Center, Pediatric Hospital, Centro Hospitalar e Universitário de Coimbra (CHUC), 3000-602 Coimbra, Portugal; Cure Sanfilippo Foundation; Dup15q Alliance; Roche Pharma Research and Early Development, Roche Innovation Center Basel, Basel, Switzerland; South London and Maudsley NHS Foundation Trust (SLaM), London SE5 8AZ, UK; Institute for Translational Neurodevelopment, Institute of Psychiatry, Psychology & Neuroscience, King’s College London, London WC2R 2LS, UK; Department of Psychiatry at The Hospital for Sick Children; Child and Youth Division Centre for Addiction and Mental Health, CAMH; Department of Psychiatry, Temerty Faculty of Medicine, University of Toronto; Génétique Humaine et Fonctions Cognitives, Institut Pasteur, UMR3571 CNRS, Université de Paris Cité, IUF, Paris, France; F. Hoffman-La Roche Ltd, Basel, Switzerland

## Abstract

**Introduction:** Autism is a common neurodevelopmental condition with a complex genetic aetiology that includes contributions from monogenic and polygenic factors. Many autistic people have unmet healthcare needs that could be served by genomics-informed research and clinical trials. The primary aim of the European Autism GEnomics Registry (EAGER) is to establish a registry of participants with a diagnosis of autism or an associated rare genetic condition who have undergone whole-genome sequencing. The registry can facilitate recruitment for future clinical trials and research studies, based on genetic, clinical, and phenotypic profiles, as well as participant preferences. The secondary aim of EAGER is to investigate the association between mental and physical health characteristics and participants’ genetic profiles.

**Methods and analysis:** EAGER is a European multisite cohort study and registry and is part of the AIMS-2-TRIALS consortium. EAGER was developed with input from the AIMS-2-TRIALS Autism Representatives and representatives from the rare genetic conditions community. 1,500 participants with a diagnosis of autism or an associated rare genetic condition will be recruited at 13 sites across 8 countries. Participants will give a blood or saliva sample for whole-genome sequencing and answer a series of online questionnaires. Participants may also consent for the study to access pre-existing clinical data. Participants will be added to the EAGER registry. Data will be shared via the Autism Sharing Initiative, a new international collaboration aiming to create a federated system for autism data sharing.

**Ethics and dissemination:** EAGER has received full ethical approval from ethics committees in the UK (REC 23/SC/0022), Germany (S-375/2023), Portugal (CE-085/2023) and Spain (HCB/2023/0038, PIC-164-22). Approvals are in the process of being obtained from committees in Italy, Sweden, Ireland, and France. Findings will be disseminated via scientific publications and conferences, but also beyond to participants and the wider community (e.g., the AIMS-2-TRIALS website, stakeholder meetings, newsletters).

**STRENGHTS AND LIMITATIONS OF THIS STUDY:**

- Data from full genotyping through whole-genome sequencing will be combined with mental and physical health data and participant research priorities
- The EAGER sample (n=1,500), although relatively small for genetic analyses, will include a substantial proportion (around one third) of participants with a rare genetic condition, ensuring that heterogeneous presentations across the autism spectrum are captured
- The EAGER registry will improve the speed, efficiency, and impact of research studies and clinical trials across Europe with a culturally diverse cohort of re-contactable participants, and shared data through the Autism Sharing Initiative
- EAGER was developed with input from the AIMS-2-TRIALS Autism Representatives and representatives from the rare genetic conditions community
- Phenotypic data are collected only via self/informant-report questionnaires and not direct clinical assessments

## 1. INTRODUCTION

Autism is a neurodevelopmental condition diagnosed on the basis of differences in social communication and interaction, sensory processing, restricted and repetitive behaviours, and intense interests (1,2), with a prevalence of around 1-2% of the general population (3). Autism is associated with a very large number of common genetic variants of small effect (4), as well as being associated with several rare genetic conditions such as Dup15q syndrome, Rett syndrome, tuberous sclerosis complex, and Angelman syndrome, where autism prevalence is as high as 30-93% (5,6).

Many autistic people have co-occurring mental and physical health conditions that impact quality of life, such as depression, anxiety, gastrointestinal symptoms, and epilepsy (7–12). For some autistic people and those with associated rare genetic conditions, many of these conditions can be severe and life-limiting, including respiratory and cardiac disorders in Rett syndrome (13,14), hyperphagia in Prader-Willi syndrome (15), and tumour growth in tuberous sclerosis complex (16). This makes the development of treatments and support for these populations a crucial area of research. Further, some autistic people may find aspects of being autistic distressing and disabling and may want help with these (17–20). All autistic people have the right to evidence-based care and support for aspects that are detrimental to their quality of life and wellbeing (21). Despite this, there is currently a paucity of personalised evidence-based care and support for autistic people across the spectrum (22,23), and polypharmacy (i.e., taking multiple medications) for various co-occurring symptoms is commonplace (24).

With a heritability of around 80%, autism has strong genetic influences (e.g., 25,26). However, to date, there is no known gene that, when mutated, increases the likelihood of an autism diagnosis without also increasing the likelihood of intellectual disability or other neurodevelopmental conditions (27). Consequently, genetic research is an important biomedical tool in the study of autism and associated rare genetic conditions, with benefits including personalised medicine, early identification of health risks, a better self-understanding, and a sense of community (28). To aid the development of evidence-based care for autistic people and those with associated rare genetic conditions, it is crucial to better understand genetic differences between syndromic forms of autism and autism without a known highly penetrant genetic influence. Further, by linking genetic mechanisms to treatment outcomes, autistic people and those with associated rare genetic conditions can be offered accurate information about what difficulties they may face both presently and in the future, and what interventions may be effective for aspects of their lived experience that they want or need support with.

EAGER (European Autism GEnomics Registry) is a European multicentre study that aims to create a registry of 1,500 participants with a diagnosis of autism or a rare genetic condition associated with autism. Participants will give a biosample and whole-genome sequencing will be performed on extracted DNA; participants will also answer a series of questionnaires related to autism, quality of life, mental and physical health, and research preferences. Using extensive genetic and phenotypic data, EAGER will enable a better understanding of the genetic architecture of autism and associated rare genetic conditions, as well as the relationship between genetics and outcomes such as health, quality of life, and wellbeing.

The EAGER registry will allow participants to be recontacted for future research studies or clinical trials that may be suitable for them. This will facilitate research by streamlining and accelerating recruitment across Europe, as well as bring together key research and clinical institutions, to the benefit of participants and their families. In line with tailoring evidence-based care to specific populations, clinical trials that are genetically-informed allow researchers to invite participants who are the most suited to, and may benefit most from, a treatment (29). Selective recruitment at the early stages of treatment development encourages the downstream development of personalised medicine.

This project aims to be a catalyst for future research and clinical trials in autism and associated rare genetic conditions in Europe. This will be achieved by connecting key sites and allowing research groups and pharmaceutical companies planning research studies or clinical trials to have access to a European registry of people who have consented to being recontacted. EAGER data will also be shared via the Autism Sharing Initiative (ASI), a global collaboration connecting autism data through federated mechanisms (30). The genetic, phenotypic, and clinical data collected in EAGER can therefore be analysed with other international autism datasets such as MSSNG (31), POND (32), LEAP (33), SPARK (34), and Searchlight (35), providing the opportunity to investigate key research questions.

## 2. METHODS AND ANALYSIS

### 2.1. Study design and population

EAGER is an international, multisite registry of adults and children with a diagnosis of autism or associated rare genetic conditions. Recruitment for EAGER will take place at 13 sites across 8 European countries (see figure 1).

**Figure 1.**
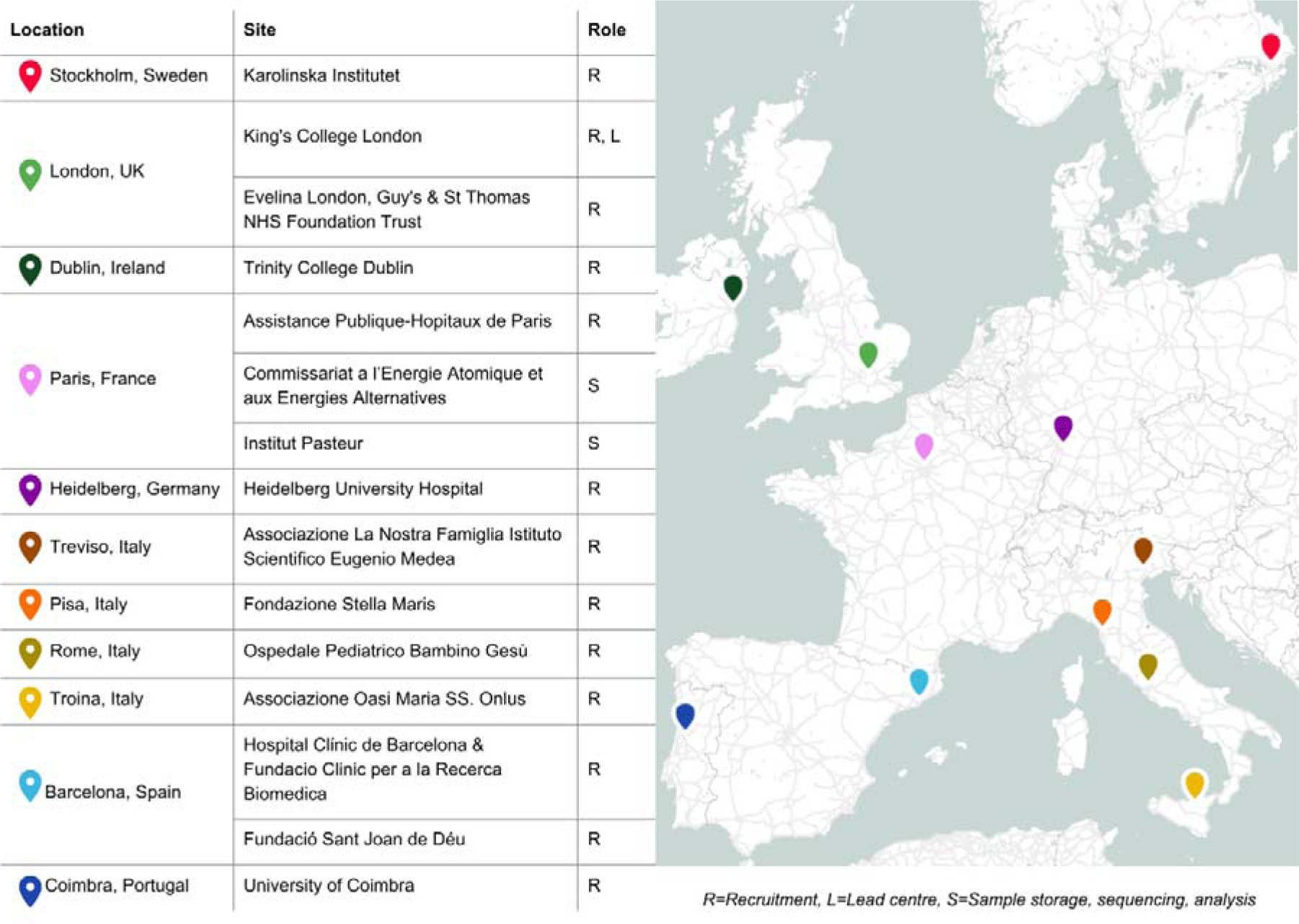
EAGER collaborating centres.

To be eligible, participants need to be over 2 years of age and have a diagnosed rare genetic condition associated with autism and/or have a diagnosis of autism (see figure 2). Decisions regarding which conditions and genetic variants to include have been made on the basis of published literature (36) and discussions with experts in the field (TB, LG), with consideration given to the penetrance, prevalence, and association with autism. For participants with a diagnosis of autism only, and where existing genetic information is available, recruitment will prioritise individuals within multiplex families and individuals with genetic variants associated with autism (see figure 2). Participants will not be excluded from the study based on any other characteristics such as co-occurring conditions, sex, or intellectual disability.

**Figure 2.**
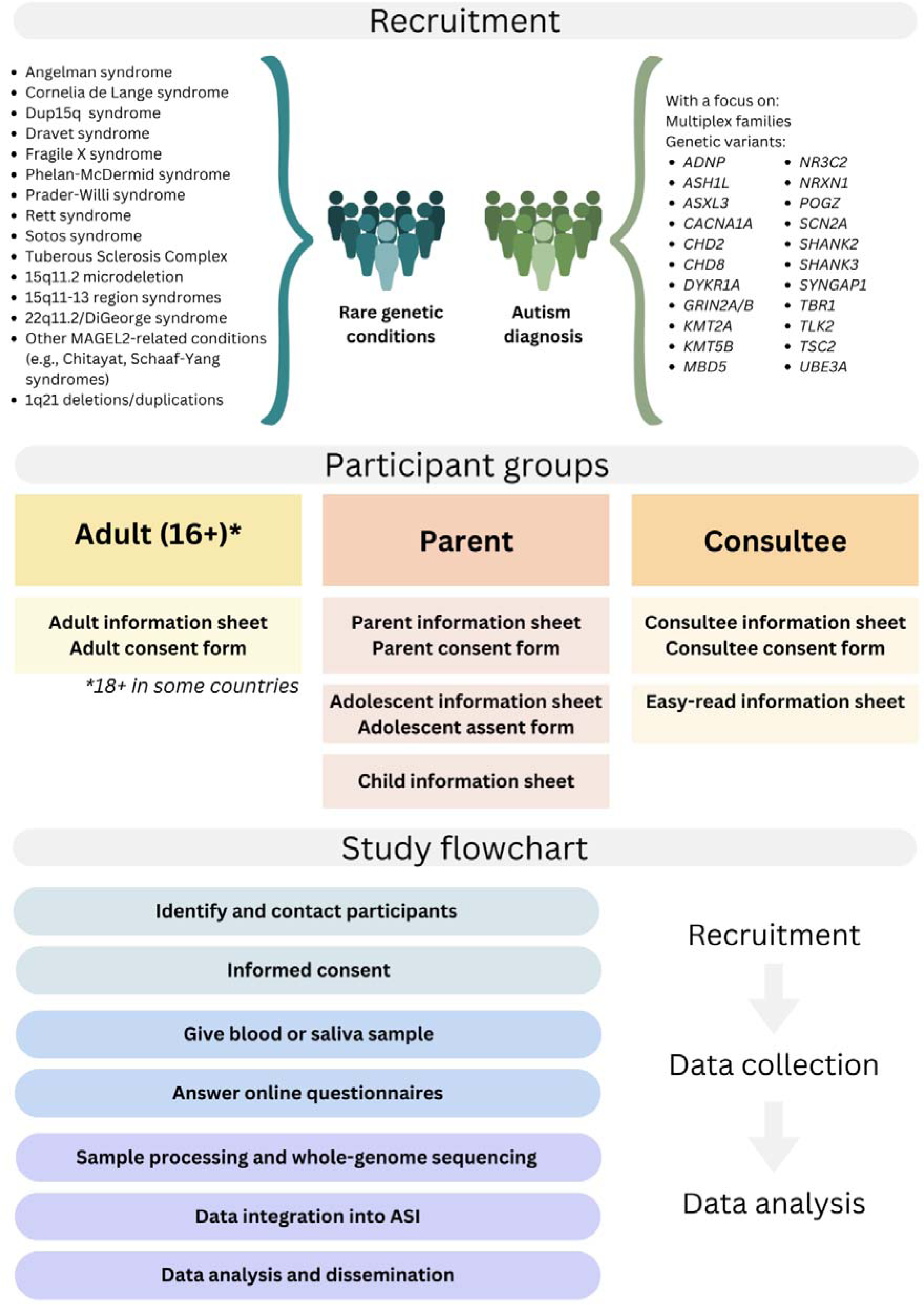
EAGER protocol summary.

### 2.2. Patient and public involvement and ethical consultations

Genetic research can involve complex ethical challenges, making it crucial to prioritise ethical considerations and participatory research practices (21,28). Participatory research practices can help ensure that the interests of our participants are appropriately represented and that the study is beneficial to its participants and the wider community. EAGER was developed with crucial input and feedback from community representatives via dedicated working groups. Our autism working group consists of AIMS-2-TRIALS Autism Representatives: autistic people and family members or carers of autistic people. Our rare genetic conditions working group consists of parents/carers of people with rare genetic conditions and/or representatives from European rare genetic conditions associations. As EAGER was conceptualised and funded prior to the establishment of our lived experience working groups, it is not a fully participatory research endeavour. This was discussed with and understood by all members of the working groups at the outset of our collaboration. Within these constraints, every effort was made to consult with our lived experience partners throughout the lifespan of this project, and feedback was integrated to the greatest degree possible.

The EAGER working groups have offered invaluable advice and feedback to guide the planning process of EAGER (see Box 1), either as part of working group meetings or through communication via email or contributions to collaborative documents. Open discussion within these groups was encouraged; as the groups included a variety of perspectives, there were naturally different solutions to questions. When no consensus was reached, the core research team made the final decision based on feasibility.

##### Box 1. Key examples of how patient and public involvement guided the EAGER study

- Refining the study rationale and clarifying perceived benefit of the study for autistic people
- Deciding on study name and logo
- Discussing ethical implications of the research, ensuring that appropriate safeguards are in place and that any limitations are transparently communicated to participants
- Ensuring that the information presented in study documents is clear, accessible, transparent, and sufficient to allow participants to make an informed decision about their participation
- Creating a “Data sharing Frequently Asked Questions” document to accompany the information sheet
- Developing different versions of information sheets tailored to the needs of our different participant groups (e.g., easy-read, adolescent information sheet)
- Ensuring that the consent forms are clear and transparent about what precisely a participant is consenting to
- Ensuring that the language used in study documents is appropriate and in line with community preferences
- Discussing the phenotypic measures used in the study and suggesting changes to the content/language of questionnaires
- Sharing all key study documents for feedback prior to submission for ethical approval
- Clearly stating that EAGER is against eugenics / prenatal testing for autism on participant-facing documents
- Detailing exactly what analyses will be performed on participant data and making it clear that no further analyses will occur without reconsenting
- Clarifying that analyses performed on EAGER data will need to be approved by a committee before they can begin
- Clarifying in the ethics documents that not all decisions were made based on consensus
- Adding a disclaimer in the protocol/ethics with regards to the fact that citing papers does not represent an endorsement of the language/aims of those papers or of the authors

Further, we conducted consultation with other stakeholders, ethical advisors, and additional working groups within the consortium, in order to evaluate risks and potentially sensitive implications of this research. We hope that the aforementioned groups will guide EAGER in the most beneficial direction for its participants and the wider community. These groups will continue to be consulted throughout the lifetime of the project.

### 2.3. Participant recruitment, information sheets, and informed consent

Potential participants will be approached by representatives at each local site. Each site will screen and recruit participants based upon recontact of participants from prior research studies or prior/current patients that fit the inclusion criteria.

Written informed consent will be required for participation in the study and participants will be provided with information sheets tailored to their age and levels of intellectual disability (see figure 2 for details on documents for specific participant groups). A personal consultee will be asked to consent on behalf of individuals who are deemed to not have capacity to consent following a capacity to consent assessment. Due to the high prevalence of co-occurring intellectual disability in people with rare genetic conditions, these may make up a significant proportion of the overall sample. Parental consent will be required for those under the legal age of consent (e.g., 16 years old in the UK).

To take part in EAGER, participants must consent to core aspects of the study: giving a blood or saliva sample (or permission to use a pre-existing sample), completing questionnaires, being added to the registry, and data sharing.

### 2.4. Biological sample collection

Participants will give a blood or saliva sample, depending on preference and appropriateness. The Oragene^®^•DNA self-collection kits, as well as assisted collection kits for those unable to collect their own sample, will be used for saliva collection. If existing blood or saliva samples are available and have been stored appropriately, they can be used in lieu of a new sample, with participant consent.

### 2.5. Measures

Following collection of the biological sample, participants will be given access to an online form with a series of questionnaires, focussing on autism, rare genetic conditions, co-occurring physical and mental health conditions, opinions on research priorities, and quality of life and wellbeing (see table 1). These will be a mixture of validated and non-validated questionnaires that have been adapted for EAGER, and have been chosen to overlap with other major initiatives or studies both within AIMS-2-TRIALS (37,38) and externally, such as MSSNG (31), POND (32), LEAP (33), SPARK (34), and Simon’s Searchlight (35). All questionnaires have been translated into the local language of each site from English. The online questionnaires will be completed by participants answering on behalf of themselves, or by parents and consultees answering on behalf of the participant.

**Table 1.** EAGER questionnaires.

| Domain | Measure | Description | Respondents |
| --- | --- | --- | --- |
| **Autism traits** | Social Responsiveness Scale (SRS (39)) | 65 items: Autism traits (including social awareness, cognition, communication, motivation, and repetitive interests and behaviours) | Adult self-report |
|  | Social Communication Questionnaire (SCQ, (40)) | 40 items: Autism traits (including communication skills and social functioning) | Parent or consultee report for child or adult |
| **Mental health** | Strengths and Difficulties Questionnaire (SDQ, (41)) | 25 items: General mental health across five scales (conduct problems, hyperactivity, emotional problems, peer problems, prosocial behaviour) | Parent or consultee report for adult or child |
|  | Patient Health Questionnaire-9 (PHQ-9, (42)) | 9 items: Core features of major depressive disorder | Adult self-report |
|  | Patient Health Questionnaire-A (PHQ-A, (43)) | 9 items: Core features of major depressive disorder* | Adolescent (16 and 17 years) self-report |
|  | Generalised Anxiety Disorder-7 (GAD-7, (44)) | 7 items: Core features of generalised anxiety disorder | Adult self-report |
| **Quality of life** | World Health Organization Quality of Life-BREF (WHO-QOL-BREF, (45)) | 26 items: Quality of life across four domains (physical health, psychological health, social relationships, environment) | Adult self-report |
|  | Cantrill Ladder (46) | 1 item: Subjective quality of life (10 point scale from “worst possible life” to “best possible life”) | Adult self-report.<br>Parent or consultee report for child or adult. |
| **Mental wellbeing** | Warwick-Edinburgh Mental Wellbeing Scales (WEMWBS, | 14 items: Mental wellbeing and health | Adult self-report.<br>Parent or consultee |
|  | (47)) |  | self-report<sup>+</sup> |
| **Restrictive and repetitive behaviours** | Adult Routines inventory (ARI, (48))<br>Repetitive Behaviours Scale-Revised (RBS-R, (49)) | 55 items: Restrictive and repetitive behaviours<br>43: Assessing restrictive and repetitive behaviours | Adult self-report<br>Parent or consultee report for child or adult |
| **ADHD** | Conners' Adult ADHD Rating Scales (CAARS, (50))<br>ADHD Rating Scale (51)) | 26 items: Core features of ADHD<br>18 items: Core features of ADHD | Adult self-report<br>Parent report for child<br>Consultee report for adult^ |
| **Demographics** | Non-validated questionnaire | 16-25 items: Demographic characteristics | Adult self-report.<br>Parent or consultee report for child or adult. |
| **Co-occurring conditions** | Non-validated questionnaire (adapted from Simons Searchlight (35)) | Items assessing 168 diagnosed and self-diagnosed psychiatric, neurodevelopmental, and physical health conditions. | Adult self-report.<br>Parent or consultee report for child or adult. |
| **Medications** | Non-validated questionnaire | 1 item: Number of medications taken by participant per day | Adult self-report.<br>Parent or consultee report for child or adult. |
| **General functioning** | Non-validated questionnaire (adapted from (52)) | 7-11 items: Seizures, gross motor skills, language and communication, intellectual disability, support needs, sleep | Adult self-report.<br>Parent or consultee report for child or adult. |
| **Voice of the community** | Non-validated questionnaire (adapted from Simon | 7-13 items: Priorities for research | Adult self-report.<br>Parent or consultee |
|  | Searchlight (35)) |  | report for child or adult. |

\* Additional questions and assessment of suicidality in the original measure were removed for the purpose of this study and only the first core 9 items were included.
^ The ADHD rating scale was adapted for use in EAGER to make it appropriate for adults and remove child-specific language.
\* Administered to parents and caregivers in relation to their own mental wellbeing.

### 2.6. Data analysis

EAGER will collect genetic and phenotypic data, which can be analysed both alone and in conjunction with other data collected within AIMS-2-TRIALS and/or shared via the ASI. Research questions will be decided in collaboration with our community working groups. Examples include identifying genetic variants that are associated with aspects of the core autism phenotype or conditions such as epilepsy or major depressive disorder, and identifying the association between genetic variants and/or polygenic scores and traits such as quality of life, levels of cognitive ability and executive function. The sample size of this study (n=1,500), which was driven by the available budget, is moderate-to-large in the field and will be significantly enhanced via the planned data sharing and integration.

## 3. DATA SHARING AND EAGER REGISTRY

### 3.1. Data sharing

Currently, funding bodies and participant groups have identified the need for research groups to share their data to maximise the value of each research contribution, pool data to address research questions that require a larger number of participants, or to carry out meta-analyses (53). For this reason, EAGER data will be shared via established AIMS-2-TRIALS data sharing mechanisms as well as being directly implemented into the ASI as detailed below. Rigorous safeguards will be implemented for access to data via established AIMS-2-TRIALS mechanisms and via the ASI platform (30).

Researchers will not be able to access EAGER data without approval from a data access committee, which will consist of autistic and non-autistic people, researchers, and members of the rare genetic community.

#### 3.1.1. Autism Sharing Initiative (ASI)

The ASI is a new collaboration working to create the first federated, global network for sharing genomic and clinical data in autism research. The ASI is led by software company DNAStack and involves other collaborators such as Autism Speaks, Roche, and the Ontario Brain Institute (please see (54) for the full list). This is an ongoing project and new collaborators may be added.

Data sharing within the ASI will be built upon decentralised, federated data sharing principles. Data federation allows researchers to search, access, and analyse data that reside in different locations and are connected via a network, without the need for data to be moved between institutions/researchers.

Data federation allows researchers to interrogate and analyse data hosted in different locations, in compliance with data regulations and improving privacy and security. A federated data network involves connecting individual datasets, which remain in their protected local environments (i.e., at the research institution where the data was collected), across one cloud-based network. To our knowledge, the ASI carries no additional risk to participant re-identification than traditional data sharing. The ASI software is built upon security standards developed by the Global Alliance for Genomics and Health (55), and regularly updated in order to ensure that data are safeguarded.

Within the ASI, data will include genomic, multi-omic, medical, and clinical data from international autism research sources. With participant consent, EAGER data (genetic data, data from online questionnaires, and existing data held at local sites) will be shared through the ASI platform with any explicit identifiers removed.

### 3.2. EAGER registry

The EAGER registry will allow participants to be contacted for future clinical trials and research studies. Researchers viewing shared participant data will be able to contact the coordinating site (King’s College London) to request specific participant groups based upon their desired criteria without access to participant personal information. King’s College London will pass these requests onto the relevant local sites, who can send information about the study or clinical trial to participants.

## 4. ETHICS AND DISSEMINATION

### 4.1. Ethics

EAGER has received full ethical approval from ethics committees in the UK (REC 23/SC/0022), Germany (S-375/2023), Portugal (CE-085/2023) and Spain (HCB/2023/0038, PIC-164-22). Approvals are in the process of being obtained from ethics committees in Italy, Sweden, Ireland, and France. Please refer to the above section “Patient and Public Involvement and Ethics Consultation” for detail on ethics and discussions within our community working groups.

### 4.2. Dissemination

We plan to disseminate our findings widely via academic avenues, such as publications in peer-reviewed major international scientific journals and conferences. In addition, we will disseminate beyond academic avenues to ensure that our findings reach participants and the wider communities, including the AIMS-2-TRIALS website (www.aims2trials.eu), stakeholder meetings, and newsletters. As part of the overall AIMS-2-TRIALS project, we have access to a European network including researchers, clinicians, and families within the autism community. This network will also provide a forum for dissemination of research findings and training opportunities.

*Disclaimer: Citing papers does not represent an endorsement of the language used in these papers, nor of the aims of the research or researchers*.

## Data Availability

Not applicable as this is a protocol paper

## AUTHOR CONTRIBUTIONS

**Conceptualisation:** MB, AL, SH, SD, PV, RP, AG, KVdB, EE, CO, VR, JT, BO, DM, LG, TB, CC, TC. **Funding acquisition:** DM, CC, TC**. Project administration:** MB, AL, DM, LG, TB, CC. TC. **Supervision:** AL, TC. **Visualisation:** MB, AL. **Data collection**: MB, AL, MA, RB, ABH, NB, SB, PB, JB, SC, RCE, MCB, JCF, PC, AD, RD, ME, MH, NM, GM, RM, CJM, SM, VM, APR, CPS, MS, KT, CT, FV, GO, BM, LG, TC. **Writing - original draft:** MB, AL, TC. **Writing - review and editing:** All authors

